## Supplementary Information for "Combining predictive models with future change scenarios can produce credible forecasts of COVID-19 futures"

**Supporting Information**

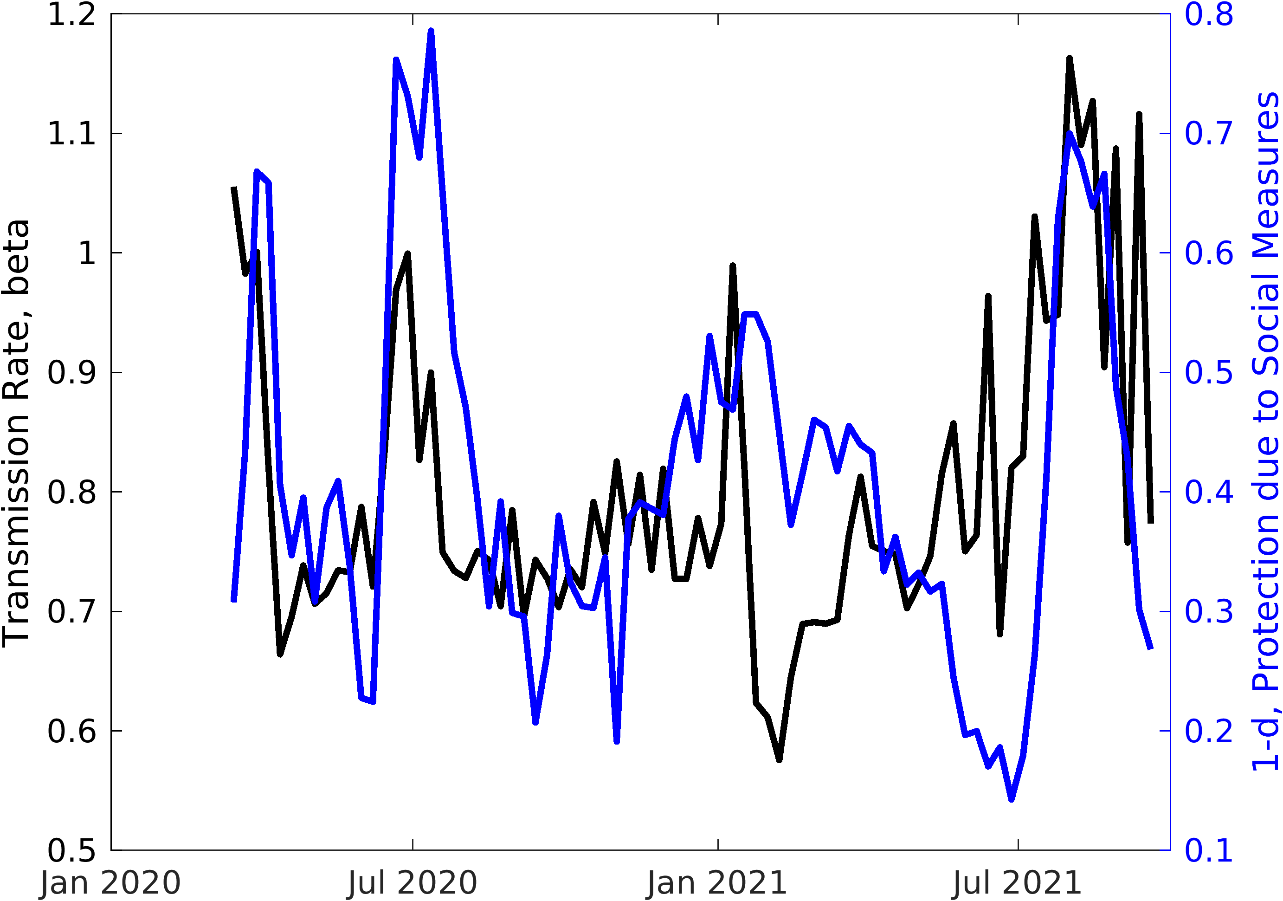

**S1 Fig.** **Average estimated transmission rate (black) and protection due to social measures (1-d parameter) over time.** The transmission rate is an averaged rate over alpha, delta, and all other variants. The priors on the d parameter are informed by Google Trends search data, as described in the main text.

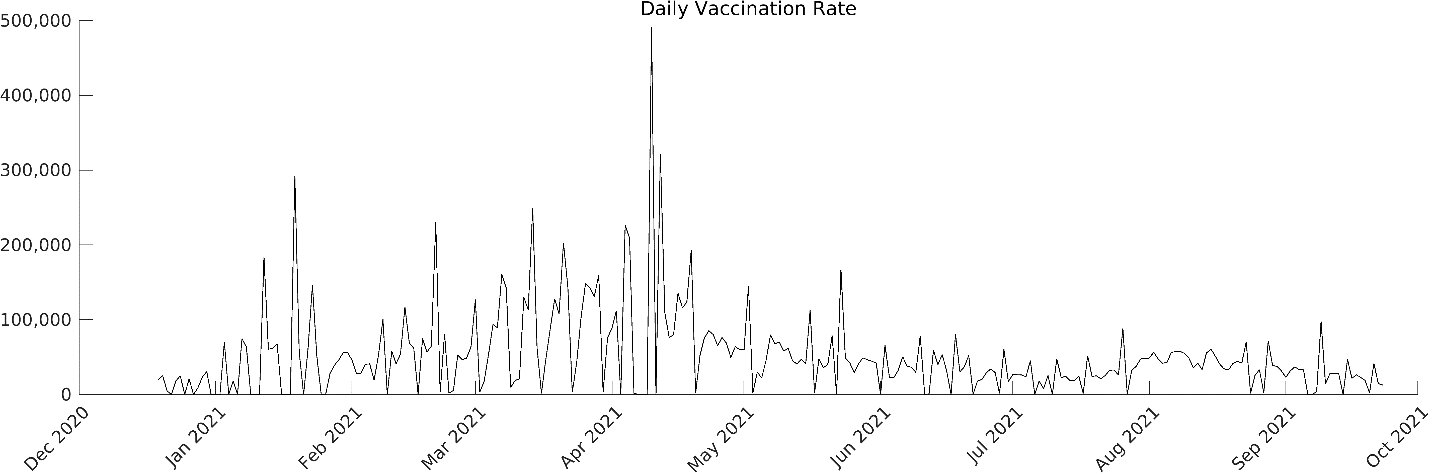

**S2 Fig. Daily reported vaccination rate in the state of Florida, as reported by coronavirus.app [60].**

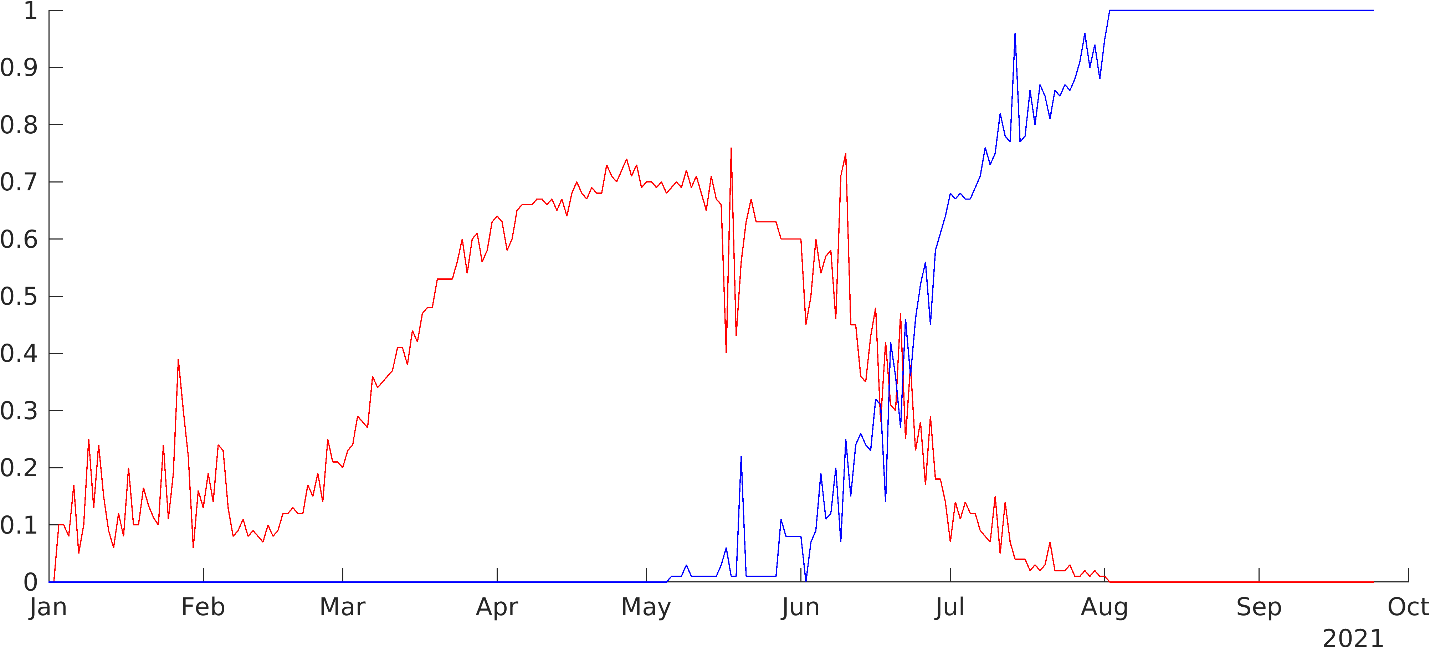

**S3 Fig. Proportion of alpha (red), delta (blue), and all other variants in the United States over time, as reported by the Helix COVID-19 Surveillance Dashboard [61]**.

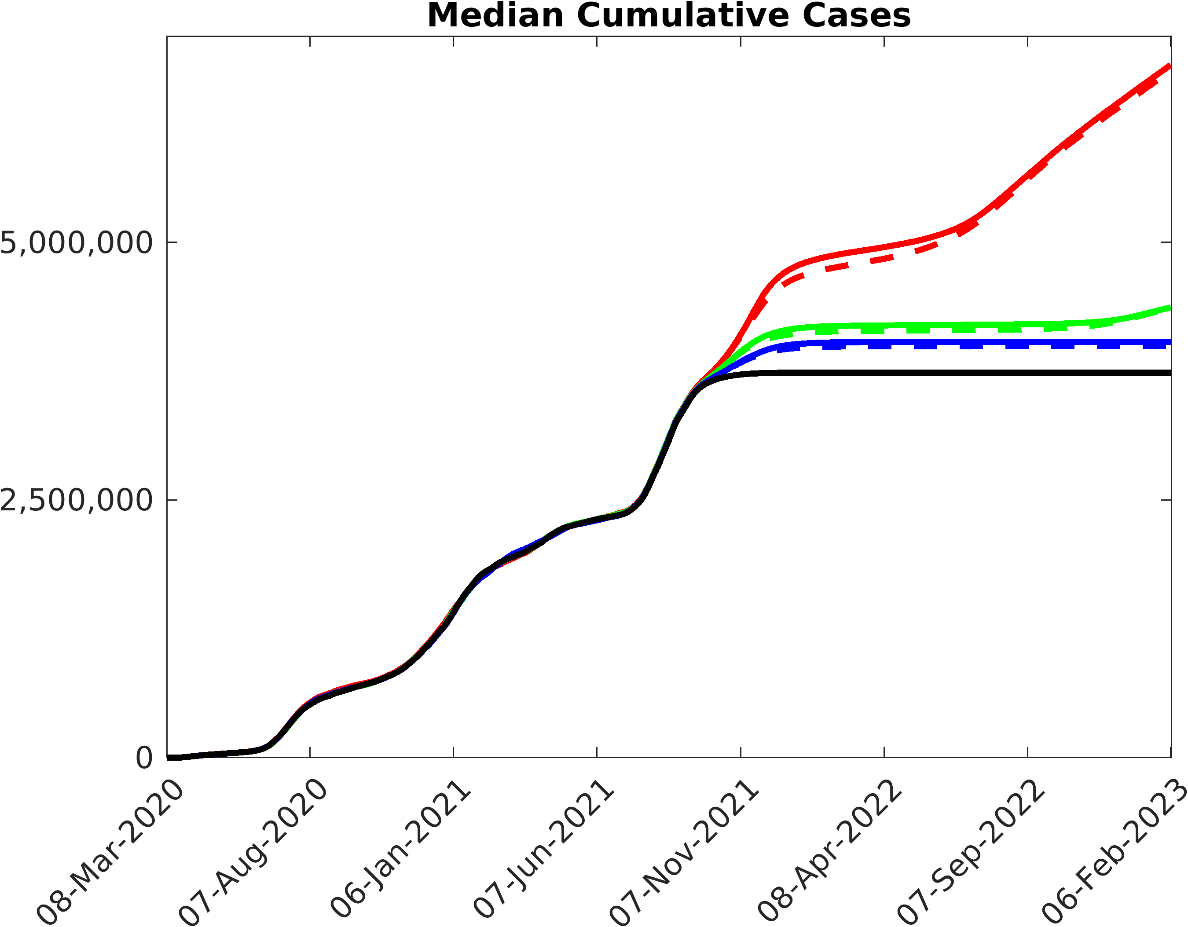

**S4 Fig. Cumulative Confirmed Cases in Florida.** The median model prediction given estimates of social measures and vaccination rate as of Sept. 24^th^, 2021 is given in black, while 1 yr, 2.5yr, and 5yr immunity waning periods are shown in red, green, and blue, respectively. The solid lines represent estimated vaccination rate, while the dashed lines represent a 1.5x increase in vaccination rate.
